# Evaluation of remote phenotyping in individuals with 3q29 deletion syndrome and development of a transdiagnostic screening protocol that can be deployed remotely

**DOI:** 10.1101/2025.10.01.25336891

**Authors:** RM Pollak, MK Harner, DV Bishop, JR Purcell, T Irving, E Sefik, C Klaiman, CA Saulnier, S Pulver, EF Walker, JF Cubells, MM Murphy, JG Mulle

## Abstract

Advances in genomics have resulted in a rapid expansion of the number of known rare genetic disorders (RGDs). However, the low frequency of RGDs presents a challenge for accurately describing the phenotypic spectrum of a given disorder. Remote phenotyping strategies are uniquely poised to address this knowledge gap. Here, we have piloted remote evaluation of cognitive ability and psychosis spectrum symptoms in 21 individuals with 3q29 deletion syndrome (3q29del) (57% male, mean age=14.3±8.6 years), a hallmark RGD. We find that remote cognitive assessment using the Penn Computerized Neurocognitive Battery and the Peabody Picture Vocabulary Test accurately captured full scale (r=0.710, p=0.001) and verbal IQ (r=0.637, p=0.003), respectively, as compared to in-person assessment with gold-standard instruments. Psychosis spectrum symptoms measured using the Structured Interview for Prodromal Syndromes were significantly correlated between in-person and remote evaluations (total score r=0.753, p=0.003; positive domain score r=0.806, p=0.0009). Based on the successful pilot of remote phenotyping in 3q29del, we designed a protocol for remote phenotyping of individuals with 3q29del. The phenotyping battery is comprised of caregiver-report and direct assessments to capture the spectrum of neurodevelopmental, neuropsychiatric, and medical features associated with the 3q29 deletion. While we designed the battery based on specific areas of concern for 3q29del, the high degree of phenotypic overlap between 3q29del and other RGDs renders this protocol amenable for implementation across a variety of RGDs, facilitating deeper understanding of the phenotypic spectrum and cross-disorder comparison. Ultimately, we hope that the increased utilization of remote phenotyping strategies will help to expand our understanding of RGDs at large, which will lead to improved clinical management strategies and better long-term outcomes for affected individuals and their families.

## Introduction

Rapid advancements in genomic technologies have led to the discovery of an increasing number of rare genetic disorders (RGDs) associated with neuropsychiatric and medical outcomes [1–4]. RGDs include single gene disorders as well as recurrent copy number variants (CNVs), caused by the deletion or duplication of large sections of DNA. It is critical to understand the phenotypic spectrum of a given RGD so that clinicians and their patients are empowered to seek optimal treatment and management strategies. Despite this need, phenotyping efforts for RGDs are fraught with challenges. The low population prevalence of RGDs is a barrier to phenotypic description [5], as individual clinicians will rarely encounter a critical number of cases in a single clinic [6]. Aggregate data from clinical case reports may provide valuable information about clinical manifestations [7–10]; however, phenotyping in such reports is rarely systematically applied, and ascertainment and publication bias may lead to only the most severe cases being reported [11]. Large, de-identified case-control studies of specific disorders such as schizophrenia can provide strong statistical evidence for associations between CNVs and specific diagnoses [12–14]. However, these studies lack data on the broader phenotypic spectrum, the prevalence of comorbidity, or the impact of the variant on individuals without a given psychiatric diagnosis. Specialty clinics or research centers allow for in-person standardized evaluations conducted by trained clinicians and provide valuable high-quality phenotypic data [15, 16], but these are only available for select RGDs, come with high operating costs, and impose a significant burden for participants, including costs for flights, hotel, and time off from work and school.

3q29 deletion syndrome (3q29del) is an RGD caused by a recurrent 1.6 Mb deletion on the long arm of chromosome 3 (hg19, chr3:195725000-197350000) [17–19]. The 3q29 deletion is extremely rare, with an estimated population prevalence of 1:30,000 individuals [5, 20]; therefore, no single clinical site sees enough patients to develop a comprehensive description of the syndrome. Case reports have documented a high prevalence of neuropsychiatric and developmental disorders, including ASD, anxiety, and schizophrenia, as well as medical challenges including heart defects and failure to thrive [7, 17–19]. Large cohort studies of people with schizophrenia have reinforced the association between 3q29del and schizophrenia, with the 3q29 deletion conferring an estimated >40-fold increased risk of schizophrenia [12, 14, 21–23]. To gain a more complete understanding of the phenotypic spectrum of 3q29del, our team previously performed in-person deep phenotyping in a cohort of 32 study participants [15, 24]. While the insights from our prior study were impactful, we were only able to describe the most common syndromic manifestations. In addition, despite our best attempts to recruit a representative sample, we cannot rule out that the burden of travel introduced a selection bias in our study.

The COVID-19 pandemic served to accelerate the development and implementation of telemedicine and remote research strategies [25–27] that are ideally suited to address the inherent challenges of phenotyping RGDs. Recent studies of multiple RGDs highlight the feasibility of remote phenotyping strategies in largely pediatric cohorts with co-occurring developmental delay and intellectual disability [28–30]. A separate study of the Bio*Me* biobank successfully administered remote clinical and cognitive assessments to adults with schizophrenia or a neurodevelopmental CNV [31]. Remote assessment is also gaining popularity in the evaluation of children with idiopathic ASD, with studies showing high concordance between remotely and in-person assessments [26, 27]. Together, these data highlight the validity of remote assessment and their utility in accessing large cohorts of patients with a rare disease. However, it is unknown whether these strategies will be effective in 3q29del. Here, we report a validated, comprehensive remote phenotyping protocol designed to capture the full phenotypic spectrum of 3q29del.

## Methods

### Prior clinical evaluation

In our prior in-person phenotyping study, individuals with 3q29del came to Emory University and were evaluated over two days using a transdiagnostic deep-phenotyping battery with gold-standard instruments administered by an expert clinical team. Anxiety symptoms were assessed using the Anxiety Disorders Interview Schedule for DSM-IV (ADIS-IV) [32] in subjects under 18 years of age or the Kiddie Schedule for Affective Disorders and Schizophrenia (K-SADS) [33] or Structured Clinical Interview for DSM-V – Research Version (SCID-5-RV) [34] in subjects 18 years of age or older. ASD symptoms were assessed in all subjects using the Autism Diagnostic Interview-Revised (ADI-R) [35] and the Autism Diagnostic Observation Schedule, 2^nd^ ed. (DAS-II) [36]. Adaptive ability was assessed in all subjects using the Vineland Adaptive Behavior Scales, 3^rd^ ed, Parent/Caregiver Form [37]. Cognitive ability was assessed using the Differential Ability Scales, 2^nd^ ed. (DAS-II) [38] in subjects under 18 years of age or the Wechsler Abbreviated Scale of Intelligence, 2^nd^ ed. (WASI-II) [39] in subjects 18 years of age and older. Executive function was assessed using the Behavior Rating Inventory of Executive Function, 2^nd^ ed. (BRIEF-2) [40] in subjects 18 years of age and younger or the Adult Version (BRIEF-A) [41] in subjects over 18 years of age. Visual-motor integration ability was measured in all participants using the Beery-Buktenica Test of Visual-Motor Integration, 6^th^ ed. (VMI-6) [42]. Prodromal symptoms and psychosis were assessed using the Structured Interview for Psychosis-Risk Syndromes (SIPS) [43] pre-interview in all subjects; subjects 8 years of age and older were also evaluated with the SIPS [43], and subjects 18 years of age and older were evaluated with the SIPS [43] and the SCID-5-RV [34]. General psychopathology was assessed using the K-SADS [33] Cross Cutting Survey and Pre-Interview in all participants; subjects 21 years of age and younger were also administered the Child and Parent Interviews; subjects older than 21 years of age were administered the SCID-5-RV [34]. Medical history, anthropomorphic measures, a medical exam, MRI imaging, and family demographics were also collected for each participant.

### Participants

To evaluate the feasibility of remote phenotyping in individuals with 3q29del, we conducted an abbreviated remote battery with individuals who had also participated in our in-person phenotyping study [15, 24]. Participants from our prior study were recontacted via email and asked to participate in the remote phenotyping pilot study. Of 34 eligible participants, 21 individuals (57% male, mean age=14.3±8.6 years; 61.8% response rate) (Table 1) completed the abbreviated battery (mean time between assessments=2.0±0.9 years). The subset of original participants that completed the remote battery was representative of the original study population; there were no significant differences between the groups for sex, age, or IQ (all p>0.05, Table S1).

**Table 1.** Description of study sample that participated in remote phenotyping pilot.

|  |  | **Mean ± SD** | **Range** |
| --- | --- | --- | --- |
| Age (years) | | 14.33 ± 8.53 | 6 - 40 |
| Composite IQ | | 74.48 ± 11.05 | 43 - 96 |
|  |  | **N** | **%** |
| Sex |  |  |  |
|  | Male | 12 | 57.14% |
|  | Female | 9 | 42.86% |
| Diagnosis |  |  |  |
|  | ASD | 5 | 23.81% |
|  | Intellectual disability | 6 | 28.57% |
|  | Graphomotor weakness | 15 | 71.43% |
|  | Anxiety | 10 | 47.62% |
|  | Prodrome/Psychosis | 3 | 14.29% |
|  | ADHD | 13 | 61.90% |

### Pilot design

The pilot remote battery focused on cognitive ability, assessed using the Peabody Picture Vocabulary Test (PPVT-5) [44] and the Penn Computerized Neurobehavioral Test Battery (Penn-CNB) [45]; and psychotic symptoms, assessed using the Structured Interview for Prodromal Syndromes (SIPS) [43]. Evaluators were trained on the administration of all assessments and certified for use prior to enrolling participants.

After remote informed consent, each participant met via a HIPAA-compliant videoconferencing platform (Zoom) with a trained research assistant for three remote study visits, lasting between 30-90 minutes each. At the first remote study visit, the researcher administers the PPVT-5. At the second and third remote study visits, the researcher administers the Penn-CNB and the SIPS, respectively. A standard protocol was in place to assess suicide and self-harm risk with clinical supports in place for further assessment if needed.

### Data analysis

We treated our prior in-person assessments as gold standard values for comparison; cognitive ability was assessed using the Differential Ability Scales (DAS-II) in individuals 18 and younger or the Wechsler Abbreviated Scale of Intelligence (WASI-II) for participants over 18. Psychosis-risk symptoms were assessed using the SIPS [15, 24, 46]. To compare performance between in-person and remote phenotyping in our cohort, we used Pearson correlation tests performed in R version 4.2.2 [47]. Remote PPVT-5 was compared to in-person measures of verbal IQ, remote Penn-CNB global accuracy was compared to in-person measures of full-scale IQ, and remote SIPS results were compared to in-person SIPS evaluation. Data visualization was performed using the ggplot2 R package [48].

## Results

### Remote phenotyping protocol validation

To determine whether remote assessments accurately captured phenotypes in individuals with 3q29del, we compared the results of our remote evaluations to prior gold-standard in-person evaluations. We found that there was a significant positive correlation between each pair of measures, indicating that the remote assessments are effectively capturing phenotypic information in 3q29del study participants as compared to in-person assessment (Table 2). The tightest correlation was between the in-person and remote SIPS measures (Figure 1, Figure S1); the SIPS total score had a correlation of 0.753 between the two measures (p=0.003; Figure 1A) and the SIPS positive domain score had a correlation of 0.806 (p=0.0009; Figure 1B). Within the cognitive domain, the remote PPVT-5 and in-person verbal IQ had a correlation of 0.637 (p=0.003; Figure 2A), and the remote Penn-CNB and in-person full scale IQ had a correlation of 0.710 (p=0.001; Figure 2B). Together, these data demonstrate that remote phenotyping in 3q29del yields results consistent with gold-standard measures.

**Table 2.** Comparison of in-person and remote methods used in pilot remote phenotyping.

| **Dimension tested** | **In-person instrument** | **Remote instrument** | **Correlation** | **P value** |
| --- | --- | --- | --- | --- |
| Verbal ability | DAS/WASI | PPVT | 0.637 | 0.003 |
| Cognitive ability | DAS/WASI | Penn-CNB | 0.71 | 0.001 |
| Psychosis-risk symptoms | SIPS | SIPS | 0.753 | 0.003 |

**Figure 1.**
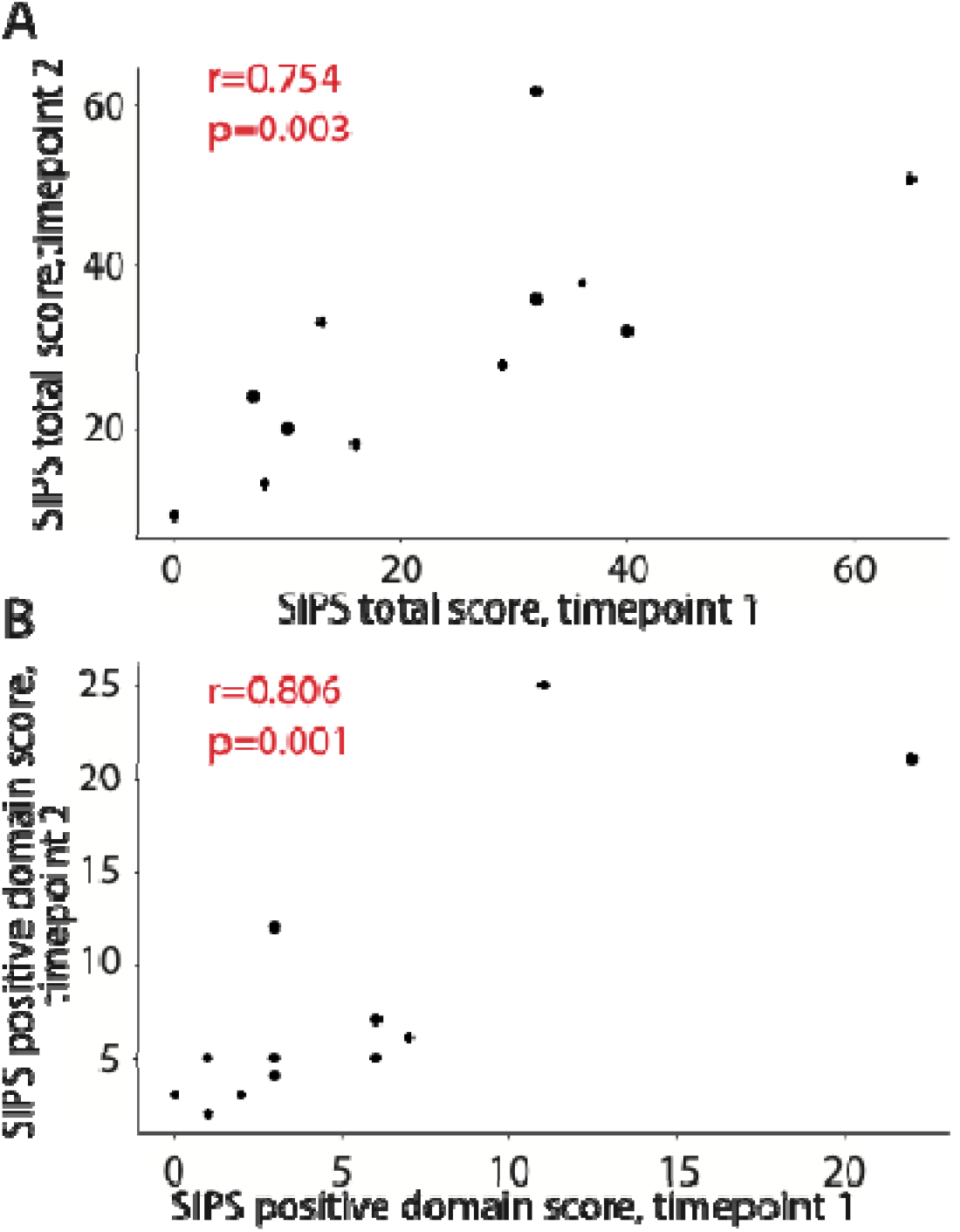
**A)** Correlation between in-person (timepoint 1) and remotely (timepoint 2) administered SIPS total score. **B)** Correlation between in-person (timepoint 1) and remotely (timepoint 2) administered SIPS positive domain score.

**Figure 2.**
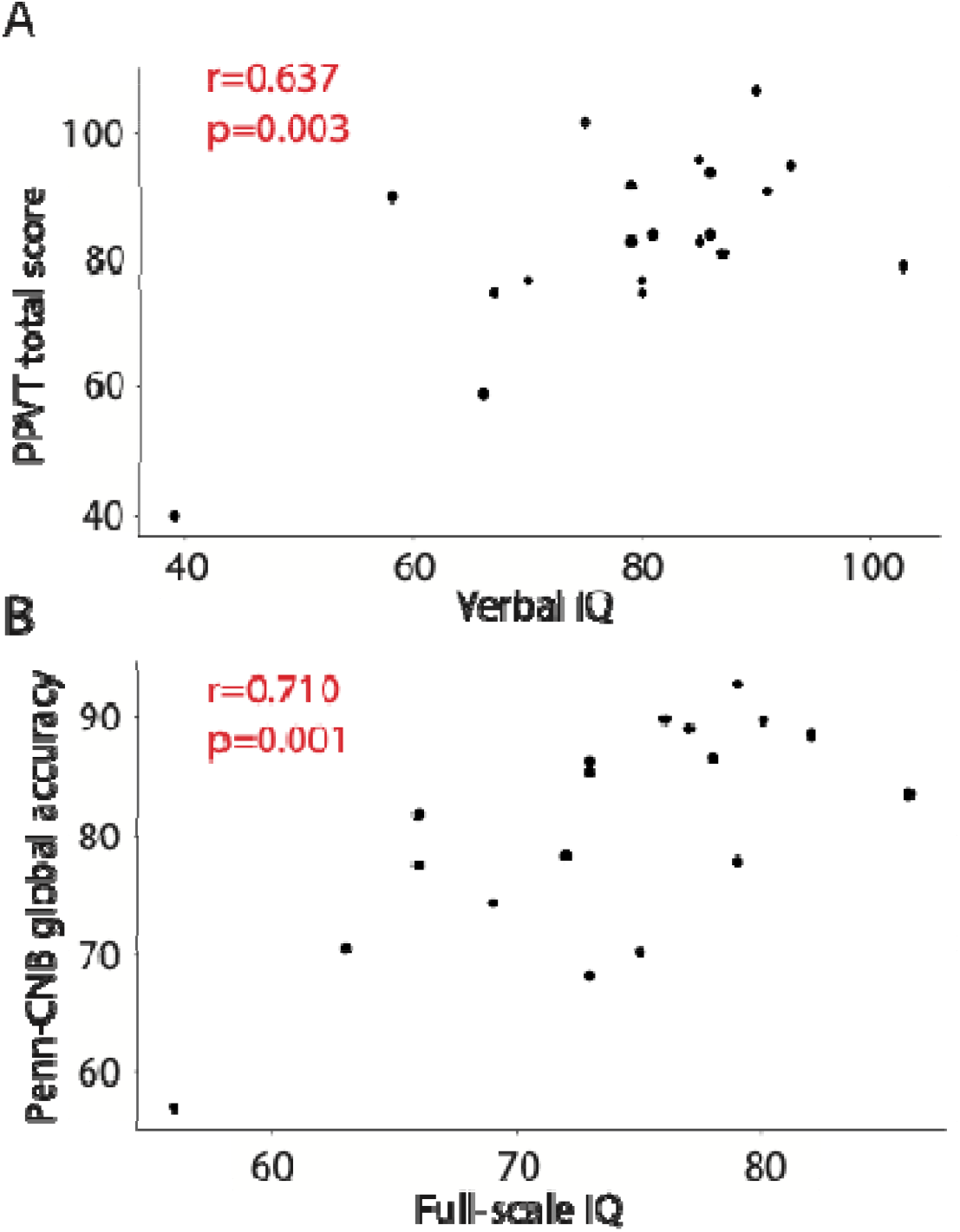
**A)** Correlation between in-person gold-standard verbal IQ and remotely administered PPVT total score. **B)** Correlation between in-person gold-standard full-scale IQ and remotely administered Penn-CNB global accuracy score.

### Development of a comprehensive remote phenotyping protocol for 3q29del

Based on the successful pilot of remote phenotyping in our study population of individuals with 3q29del, we moved to create a comprehensive remote phenotyping battery designed to measure the primary domains and subdomains that are impacted in individuals with 3q29del.

## Participants

We will recruit individuals with the 3q29 deletion from the existing online 3q29 Registry [18], as well as direct referrals and contacts. To be eligible for the study, probands must be eight years of age or older, fluent in English, and have a clinically confirmed diagnosis of 3q29del. Non-verbal participants will be administered an abbreviated study protocol. Participants and their caregivers must be willing and able to provide informed consent and be fluent in English to participate.

Exclusion criteria are non-fluency in English and age younger than eight years. As a comparison sample we will enroll typically developing controls recruited from community sources and online platforms for individuals interested in participating in research (ResearchMatch). All enrollment, consenting, and data collection procedures are identical between 3q29del probands and typically developing controls.

## Phenotyping battery

### Executive function, attention, and hyperactivity

Executive function is assessed using the Behavior Rating Inventory of Executive Function, 2^nd^ Edition (BRIEF-2) for participants 18 years of age or younger, or the adult version (BRIEF-A) for probands over 18 years of age [40, 41]. Caregivers rate behaviors associated with self-control and problem-solving skills along nine dimensions of executive functioning: inhibiting distractions, self-monitoring, shifting, emotional control, initiation, working memory, planning, organization, and task monitoring. Probands age 21 or younger are also assessed using caregiver-report on the Conners (Conners-3 or Conners-4), a questionnaire designed to assess for DSM-V symptoms of ADHD, conduct disorder, and oppositional defiant disorder [49, 50]. This age cutoff was established due to the phenotype of developmental delay frequently observed in those with 3q29del [7, 15, 18]. Probands over the age of 21 are assessed for attentional deficits via the analogous Conners’ Adult ADHD Rating Scale (CAARS or CAARS-2) [51].

### Cognitive ability and receptive vocabulary

Receptive verbal comprehension is assessed in probands at the first study visit using the Peabody Picture Vocabulary Test-5 (PPVT-5), a computerized, norm-referenced measure of receptive vocabulary based on words in Standard American English [44]. On the PPVT-5, probands hear a word and select one of four illustrations that best depicts the word. The Penn Computerized Neurocognitive Battery (Penn-CNB) is conducted with each subject as a measure of cognitive ability across five domains: executive, episodic memory, complex cognition, social cognition, and sensory/motor speed [45]. The battery consists of 14 computerized tasks and puzzles designed to measure the accuracy and speed of the participants’ responses (full list of tasks in Supplemental Material). The battery is administered remotely by a certified researcher who provides standardized instruction, ensures participant understanding, and takes detailed notes on any issues that may compromise data quality, such as software glitches, interruptions, or idiosyncratic participant behavior.

### Adaptive behavior

Adaptive behavior is assessed using the Vineland Adaptive Behavior Scales-3 (Vineland-3), a standardized caregiver-report assessment of an individual’s ability to independently perform age-appropriate day-to-day activities across four domains: communication, daily living skills, socialization, and motor skills [37].

### Social function related to ASD

Social function relevant to ASD is assessed using the Social Responsiveness Scale (SRS), which measures social information exchange across five subscales: social awareness, social cognition, social communication, social motivation, and restricted interests and repetitive behaviors [52].

Caregivers complete an informant-report form on their child’s behaviors. Additionally, caregivers complete the Social Communication Questionnaire (SCQ) as a secondary measure of social function in the proband [53]. The SCQ is derived from the Autism Diagnostic Interview-Revised (ADI-R) [35], evaluating social behavior across three domains: reciprocal social interaction; communication and language; and restricted, repetitive, and stereotyped interests and patterns of behavior.

### Psychosis risk symptoms and psychosis

The Structured Interview for Psychosis-Risk Syndromes (SIPS) assesses the presence, duration, and severity of subthreshold symptoms of psychosis [43]. The SIPS is a gold-standard semi-structured interview used to assess prodromal symptoms of psychosis. Pre-interview questions are asked at the beginning of the caregivers’ remote study visit. The SIPS interview is then conducted by certified interviewers remotely with the proband without the caregiver present, unless there is a strong preference for caregiver attendance for the proband’s comfort or intellectual disability. The SIPS is recommended to be conducted with interviewees between the ages of 10 and 45, with an IQ above 70; participants not meeting these criteria will be administered the SIPS but partitioned out of the final analysis. When necessary, corroborative information is ascertained from a caregiver in a follow-up interview to resolve questionable information reported by probands who exhibited difficulty with understanding or answering interview questions. Participants are rated on a scale of 0-6 across positive, negative, disorganized, and general symptom domains, with scores of 3 or greater on any item indicating clinically significant prodromal symptoms.

### General psychopathology

General psychopathology is assessed using the Adult Behavior Checklist (ABCL) for participants 18 years of age or older or Child Behavior Checklist (CBCL) for participants younger than 18 years of age [54, 55]. These are standardized, caregiver-report questionnaires that measure the presence of behavioral and emotional problems across eight syndrome scales: anxious/depressed, withdrawn/depressed, somatic complaints, social problems, thought problems, attention problems, rule-breaking behavior, and aggressive behavior.

### Graphomotor function

The Beery-Buktenica Developmental Test of Visual-Motor Integration – 6^th^ edition (Beery VMI) measures visual-motor integration [42], which is impaired in individuals with 3q29del (mean score=69.3±16.7) [56]. The Beery VMI requires probands to copy each of 28 abstract designs of increasing complexity. Caregivers are instructed on proper administration of the Beery VMI during their remote study visit and step-by-step written instructions are included in the study kit. Probands complete the Beery VMI with caregiver oversight outside of study visits. The worksheet is returned with the study kits and scored by the study team.

### Medical/physical

A detailed medical history questionnaire (see Supplemental Materials) is completed online by the caregiver. The survey includes questions to assess for pregnancy history, including diagnostic testing/screening, prescriptions or supplements taken, and drug or alcohol exposure; birth history, including complications, birthweight, and gestational age; developmental history, including motor and communication milestones; and a systems-based medical history. The survey has been adapted from our previous study [24]. Height, weight, and head circumference measurements are collected by caregivers using standardized equipment and procedures (see Supplemental Materials) to evaluate the physical stature of probands.

### Medication history and efficacy

A lifetime psychiatric medication history questionnaire is also completed by caregivers. A list of psychiatric medications is provided. For each medication that the proband has taken, either currently or in the past, the caregiver records dosage, duration, indication, side effects, and perceived efficacy.

### Data collection procedures

Data are collected via remote study visits, online self-report or informant-report surveys, and at-home data collection by the caregivers with instruction from trained researchers (Table 3). For adult probands, a spouse or sibling may participate as an informant in lieu of a caregiver. Figure 3 summarizes the study protocol. Once informed consent is obtained, demographic data are collected via a custom REDCap survey, and remote study visits are scheduled using a HIPAA-compliant online scheduling platform. A study kit including necessary study materials and instructions is mailed to the family (see Supplemental Material), and links to online surveys are emailed to the caregivers. Online assessments are administered through REDCap and instrument publishers’ websites. Assessments and questionnaires have been selected to measure the primary domains and subdomains that are impacted in individuals with the 3q29 deletion [15, 18].

**Table 3.**
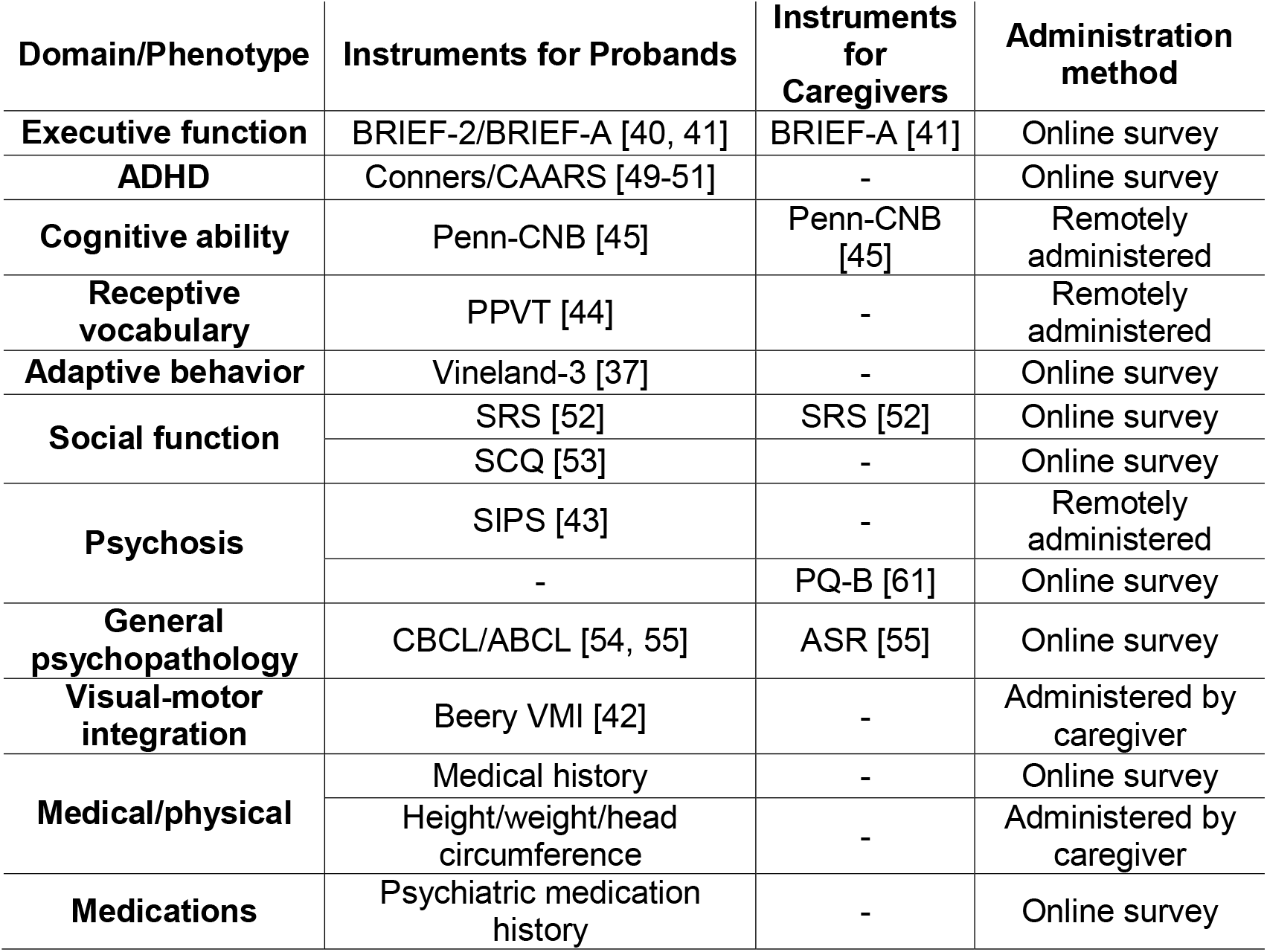
Assessments and domains measured. Complete list of assessments and surveys to be completed by probands and caregivers, categorized by domain. Online surveys are completed by caregivers. “Remote administration” assessments are administered over Zoom by a trained researcher. “Administered by caregiver” assessments are administered by a caregiver after receiving instruction from a trained researcher.

**Figure 3.**
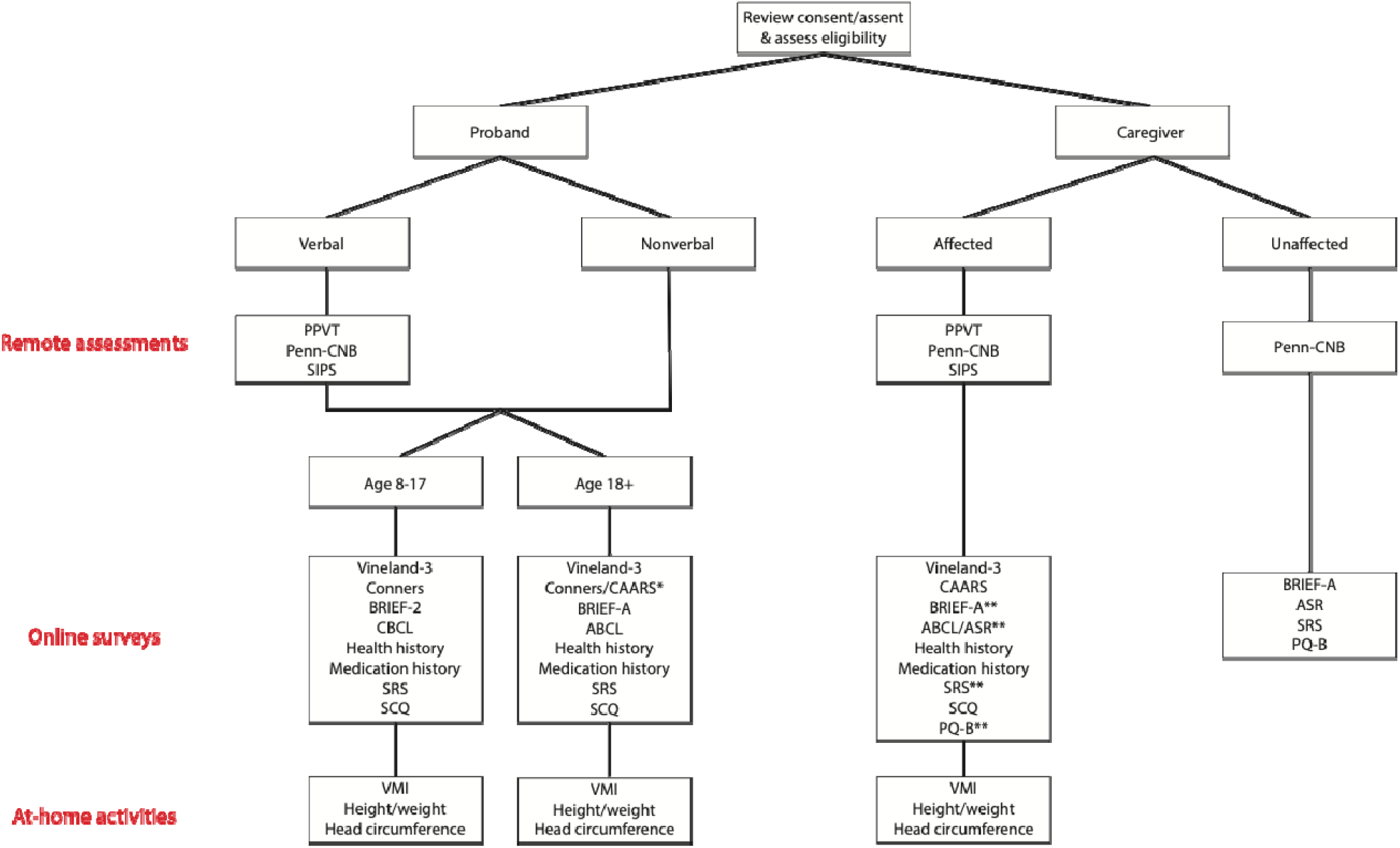
Flow chart of study design. *The Conners is administered to adult probands ages 18-21, while the CAARS is administered to probands over the age of 21. **Affected caregivers (those with the 3q29 deletion) complete the BRIEF-A, ASR, SRS, and PQ-B as self-report instruments, in addition to the standard proband battery of informant-report surveys which are completed by a parent, spouse, or other informant.

The study kit also includes materials to collect height, weight, and head circumference data on the proband (Supplemental Materials).

Probands meet via Zoom with a trained research assistant for three remote study visits, lasting between 30-90 minutes each. At the first remote study visit, the researcher administers the PPVT-5 [44] and provides instruction for measuring the proband’s head circumference using the provided materials. Head circumference is measured on-camera with guidance from the researcher to ensure an accurate measurement. At the second and third remote study visits, a researcher administers the Penn-CNB [45] and the SIPS [43] to the proband, respectively. A standard protocol is in place to assess suicide and self-harm risk with clinical supports in place for further assessment if needed. Non-verbal study participants are not required to complete the study visits. One caregiver is also queried on their child’s developmental, social, and psychological history, as well as any family history of mental illness.

Upon completion of all data collection instruments, participants return the study kit using an included pre-paid shipping label. The research team collates all questionnaire and assessment data into a written research evaluation report to be returned to each family. A follow-up videoconferencing session is then arranged between the proband/caregivers and the research team, at which point the research evaluation report is emailed to the family and results are discussed with the proband and/or their caregivers.

## Discussion

Here we have demonstrated the validity of remote phenotyping in individuals with 3q29del and present a protocol for remote evaluation of multidimensional phenotypes associated with 3q29del. By harnessing the advantages of remote evaluation, this protocol allows for the administration of standardized assessments across core domains previously found to be impacted in individuals with 3q29del. The proposed protocol allows for collection of multidimensional behavioral data at scale, while minimizing participant burden and maximizing feasibility.

Remote study designs have several important advantages over traditional in-person clinic when considering RGDs. By definition, remote assessment can be performed anywhere, eliminating the need to travel to a specialty clinic for evaluations. Travel is a significant barrier to care for individuals with RGDs, as it is costly, requires significant absences from school or work, and can be stressful for the affected individual and their family members [57]. Additionally, when an individual goes to a specialty clinic to receive care, there are typically numerous sessions and evaluations in a day, which can lead to testing fatigue. Remote studies allow the evaluations to be divided across multiple sessions, reducing the strain on the participant and ensuring that the testing can be completed at times that are convenient for the family’s existing schedule and obligations. Finally, remote study designs can serve to expand the spectrum of recruitment for a given RGD – it is possible that the most severely affected individuals are unable to travel, and higher-functioning individuals may choose not to participate in in-person studies because they do not require a high level of specialty care. Convenient, at-home remote assessments can serve the needs of the full spectrum of abilities associated with RGDs and reduce ascertainment bias.

While we specifically designed the protocol presented here for phenotyping 3q29del, it should be noted that it is also generalizable to other RGDs given the high degree of phenotypic overlap, especially among CNV disorders. Developing a consistent, standardized phenotyping battery will facilitate cross-disorder analysis [58]; without core phenotyping instruments, it is difficult to impossible to identify areas of phenotypic convergence and divergence. Indeed, some of the instruments included in the present battery have already been implemented in other disorders; namely, the Penn-CNB in 22q11.2 deletion syndrome [59] and the SIPS in the North American Prodrome Longitudinal Study of individuals at high risk for psychosis [60]. Harmonizing instruments across studies allows for the well-powered cross-disorder analyses that will ultimately lay the groundwork for studies of the mechanisms underlying phenotype development in these RGDs.

The protocol described here is an important step toward a more comprehensive understanding of 3q29del, and ideally a harmonized phenotyping approach for RGDs at large. However, there are important limitations to consider when discussing remote phenotyping. First, the researchers conducting remote phenotyping assessments have significantly less control over the testing environment and possible distractions as compared to an in-person study. It is also harder for the researcher to make granular behavioral observations, such as observations about eye contact and social cuing. To address these concerns, our researchers take detailed notes during all sessions to document any perceived distractions or interruptions that may compromise data quality.

Although remote studies can increase ascertainment across the phenotypic spectrum, individuals with poor internet connection or limited access to technology are unable to participate in remote studies. In our pilot study, we were concerned about biased ascertainment due to technology access. To protect our findings against this possibility we obtained a study laptop that could be shipped to families that needed it. To our surprise, all study participants had access to a laptop or desktop computer, regardless of socioeconomic status. This is likely a consequence of the pandemic and the rise of remote classroom strategies. Participant retention across multiple study visits is also a concern; however, we have found that families are highly motivated to complete all required assessments in order to receive their final report with the testing results. Finally, it can be difficult to accurately measure cognition remotely for the youngest and most severely affected individuals; the Penn-CNB is validated for remote use [59] but does require basic verbal comprehension, focus, and motor skills to complete the tasks. Our researchers take detailed notes during the Penn-CNB administration to assess participant understanding of the tasks, idiosyncratic behavior, and reaction time, to ensure that participants understand and are able to complete each task. To supplement the Penn-CNB, we are also collecting data on verbal ability using the PPVT-5 [44] and we request any prior cognitive testing reports that participants have. Additionally, our validation analysis shows that both the Penn-CNB and the PPVT-5 performance are strongly positively correlated with gold-standard in-person IQ measures. We have observed that when 3q29 deletion individuals are non-verbal or have lower cognitive ability such that they cannot effectively complete the PennCNB, they almost always have alternate cognitive testing results.

This is the first report of remote cognitive and behavioral phenotyping in individuals with 3q29del, as well as a description of a standardized remote testing battery for phenotyping 3q29del and phenotypically similar RGDs. It is critical to develop a comprehensive understanding of the phenotypic spectrum of RGDs in order to improve treatment plans and outcomes for affected individuals. Further, expanding our understanding of RGDs such as the 3q29 deletion may also lead to mechanistic insights about the development and pathogenesis of common psychiatric disorders such as schizophrenia and ASD. We hope to inspire other groups to adopt a standardized phenotyping protocol to facilitate deep understanding of individual RGDs as well as comparison across RGDs, which will ultimately benefit clinicians, families, and affected individuals by helping to guide care plans and improve treatment outcomes.

## Data Availability

All data produced in the present study are available upon reasonable request to the authors.

## Notes

### Competing Interest Statement

CAS reports receiving royalties for the Vineland-3.

### Funding Statement

This study was funded by the National Institute of Mental Health (MH126449).

### Author Declarations

IRB of Rutgers University gave ethical approval for this work. IRB of Emory University gave ethical approval for this work.

