## Supplemental Information for "Evaluation of remote phenotyping in individuals with 3q29 deletion syndrome and development of a transdiagnostic screening protocol that can be deployed remotely"

**Table S1.** Comparison of subjects who participated in remote pilot to subjects that did not participate.

|  |  | Remote and in-person | | In-person only | | P value |
| --- | --- | --- | --- | --- | --- | --- |
|  |  | Mean ± SD | Range | Mean ± SD | Range |  |
| Age (years) | | 14.33 ± 8.53 | 6 - 40 | 17.54 ± 7.58 | 8 - 35 | 0.263 |
| Composite IQ | | 74.48 ± 11.05 | 43 - 96 | 68.38 ± 19.09 | 40 - 99 | 0.31 |
|  |  | N | % | N | % |  |
| Sex | |  |  |  |  | 0.733 |
|  | Male | 12 | 57.14% | 9 | 69.23% |  |
|  | Female | 9 | 42.86% | 4 | 30.77% |  |

**Figure S1.** SIPS profiles at timepoint 1 (in-person evaluation) and timepoint 2 (remote assessment) for all pilot study participants.

***1. Study Kit Contents and Participant Instructions***

A Study Kit is shipped to each participating family. The Study Kit contains the materials required for the physical measurements and other study-related items. Below is a detailed breakdown of the contents of the Study Kit:

1. **Anthropometric Measurements:** Weight, height, and head circumference measurements are collected using materials and instructions included in the study kit. Instructions for taking height and weight measurements are included in the study guide (see **Figure S2**).
   a. **Body Weight:** A digital scale is provided for accurate measurement of body weight; the scale is pre-set to kilograms. Families are instructed to record two weight measurements (three if the first two weights differ) for the proband on an included data card to ensure accuracy and to reduce calibration errors which can occur during shipment.
   b. **Height:** A measuring tape is included as well as painters’ tape to secure the measuring tape to the wall. Families are instructed to mark the probands height in centimeters on the measuring tape with a permanent marker (included) and return it with the Study Kit.
   c. **Head Circumference:** A retractable measuring tape is included to measure the probands head circumference. A researcher instructs a caregiver on how to properly take and record the measurement by marking it with a permanent marker (included) on the tape, which is then returned with the Study Kit at the end of the study. Head circumference is measured on camera in the presence of a researcher to ensure an accurate measurement.
2. **Documents:** A Study Guide, signed copies of the informed consent forms, the *Beery-Buktenica Developmental Test of Visual-Motor Integration* (*VMI*) assessment packet, and a return shipping label are enclosed in an envelope within the Study Kit. Families are instructed on how to properly assess the proband with the *VMI* during the one of the caregiver study visits, and instructions are included in the Study Guide (see **Figure S3**). The Study Guide also contains a checklist of surveys and activities for each participant; a general outline of each study visit, survey, and assessment; and instruction for returning the Study Kit at the end of the study, along with the address of the nearest UPS drop-off location.

***Supplemental Figure S2: Instructions for Taking Height and Weight Measurements***

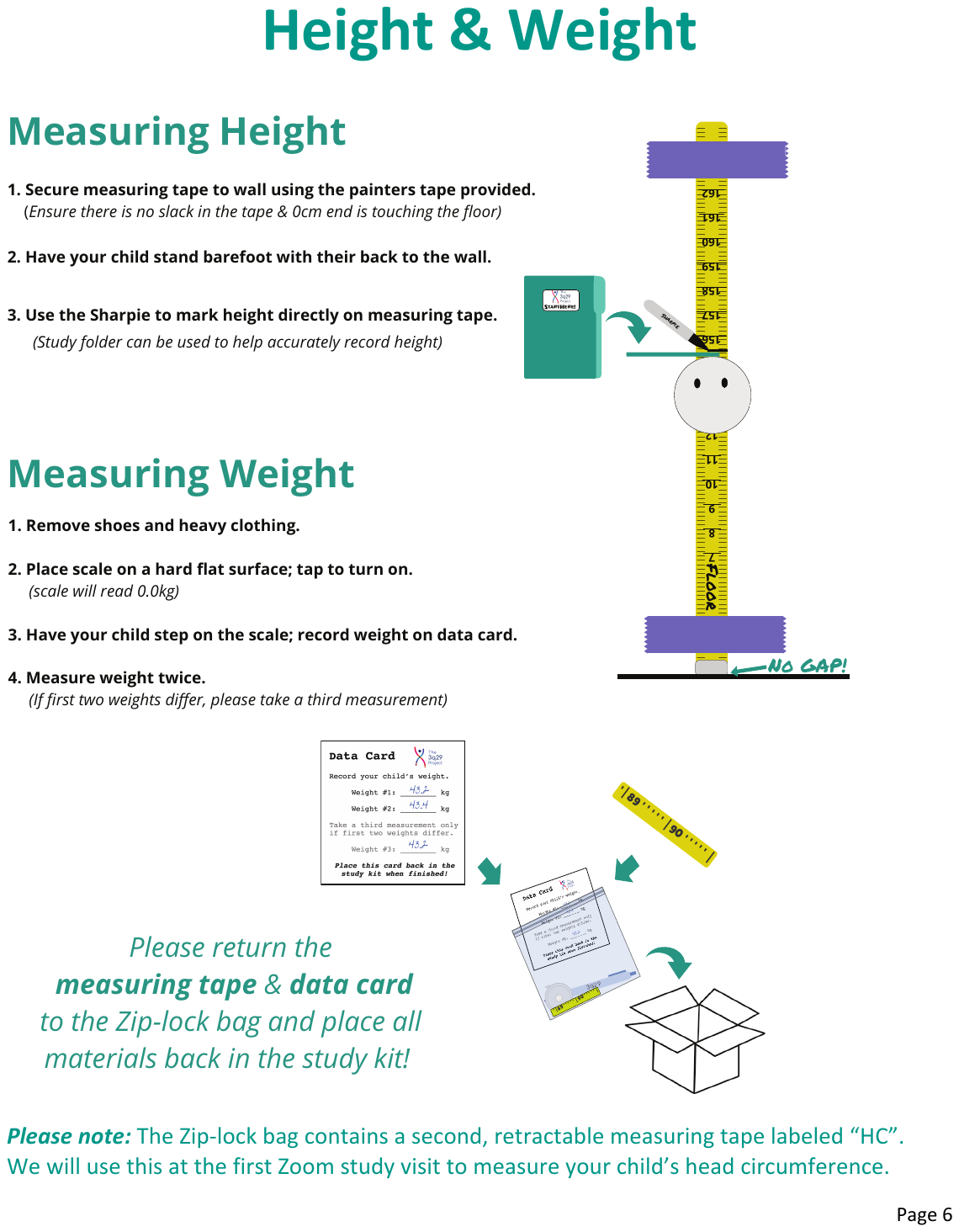

***Supplemental Figure S3: Instructions for Administering the VMI***

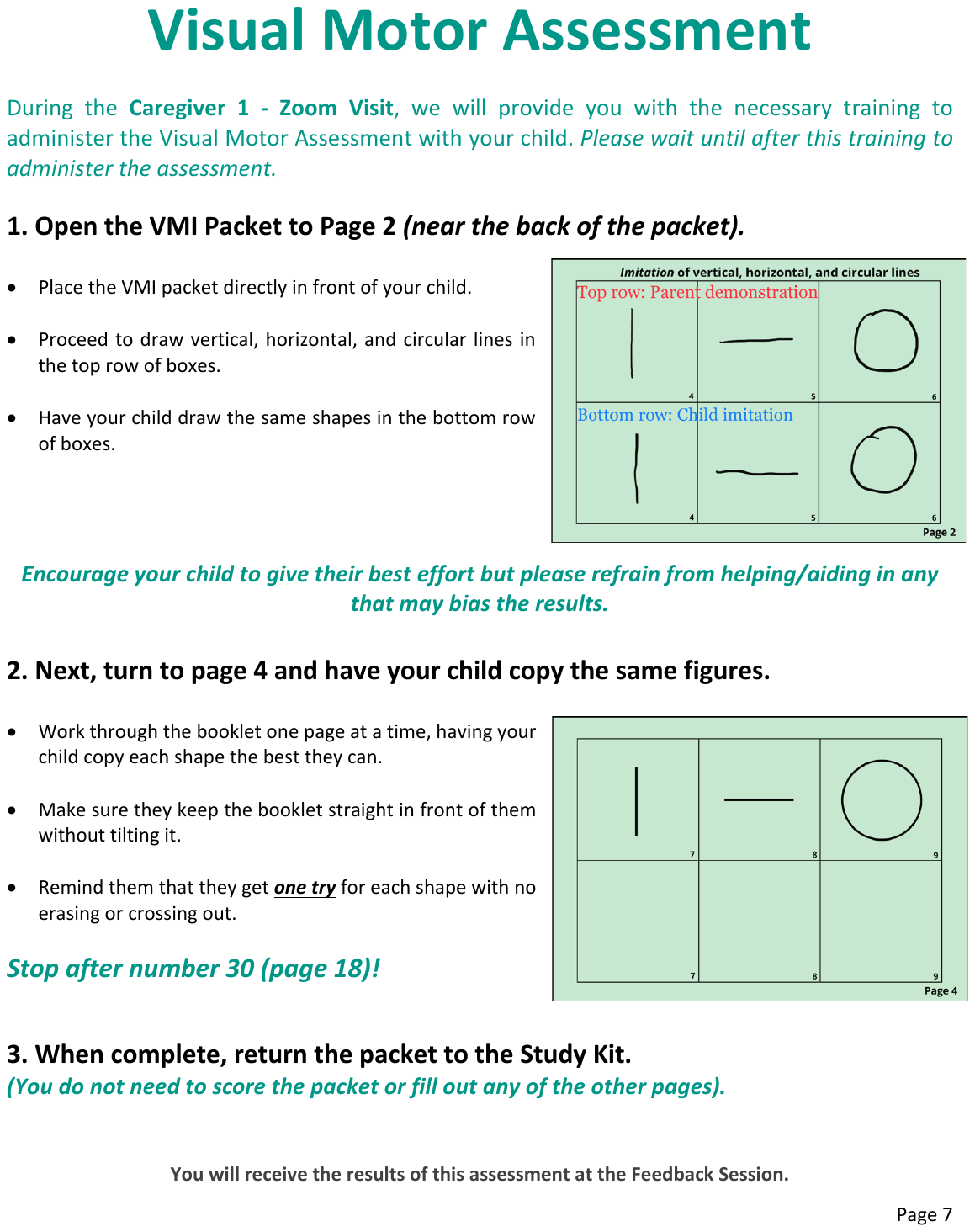

### ***2. Penn Computerized Neurobehavioral Test Battery***

As a measure of cognitive ability, this study utilizes the *Penn Computerized Neurobehavioral Test Battery (CNB)*, which consists of a series of cognitive tasks designed to measure cognitive performance across multiple domains. In the present study, the *CNB* is conducted remotely; a certified researcher administers the battery with each participant over Zoom, providing instructions, ensuring task comprehension, and noting any potential problems which may affect data quality (i.e. distractions, poor internet connection, motor issues). The subtests included in the version of the battery utilized in the present study were selected with the known profile of cognitive ability in 3q29 deletion. There are 14 subtests included in the battery, as follows:

1) Motor Praxis

2) Measured Emotion Differentiation Test

3) Verbal Reasoning Task

4) Facial Memory Test

BREAK POINT

5) Emotion Recognition Test

6) Word Memory Test

7) Age Differentiation Task

8) Variable Short Line Orientation Test

BREAK POINT

9) Visual Object Learning Test

10) Matrix Analysis Test

11) Short Continuous Performance Test

BREAK POINT

12) Conditional Exclusion Task

13) Go No Go Task

14) Short Computerized Finger Tapping Test

***3. Health History Survey***

Information on pregnancy, birth, developmental, and medical histories are queried using a caregiver-report survey on REDCap. There is an option to upload prior cognitive testing results at the bottom of the survey.

***Survey Questions:***

- Please answer the following questions about the individual with 3q29 deletion.
  - When did you or your primary care provider first suspect a problem?
  - When was your child first diagnosed with 3q29 deletion?
  - What current questions or concerns do you have about your child?
  - Is your child adopted?
- The following section asks about the individual's biological (birth) parents. If you know information about the birth parents, pregnancy, and delivery, please answer the questions accordingly; otherwise, you may leave these questions blank.
- Pregnancy History (for the pregnancy of the person with 3q29)
  - Mother's age (in years) at delivery?
  - Fathers's age (in years) at delivery?
  - What number pregnancy was this for the mother?
- Please answer the following Yes/No questions about the pregnancy. If yes, please provide detail below.
  - Prenatal vitamins?
  - Medications (prescription)?
  - Medications (over-the-counter)?
  - Smoking?
  - Alcohol (beer, liquor, wine)?
  - Street drugs?
  - Illness/infection?
  - Bleeding?
  - Rash?
  - Fever?
  - Diabetes?
  - High blood presure?
  - Thyroid problems?
  - X-ray/radiation?
  - Premature labor?
  - Hospitalization (do not count the delivery/birth)?
  - Abnormal growth of baby?
  - Other concerns?
- Please answer the following Yes/No questions regarding testing that may have been done during the pregnancy.
  - First Trimester Screen (ultrasound of baby's neck/Nucal Translucency/NT measurement plus blood work)
  - Second Trimester Screen (Triple Screen, Quad Screen, AFP Test)
  - Chronic Villus Sampling (CVS)
  - Amniocentesis
  - Glucose Tolerance Test
  - Routine Ultrasound
  - Specialized Ultrasound
  - Other Testing. If yes, Please explain other testing that may have been done during the pregnancy:
  - Were any of the screening, diagnostic, or other tests ABNORMAL? If YES, please explain:
- Birth History (for the person with 3q29)
  - Due date:
  - Date delivered:
  - How was the child delivered? If C-section, Please explain the reason why a C-section was performed (i.e. previous child born that way, failure to progress, etc.):
    - Vaginal
    - C-section
  - Was the baby born head first?
  - Baby's weight (in pounds):
  - Were there complications with the delivery? If yes, Please list complications with the delivery:
  - Were there any problems right after birth (i.e. need to go to the NICU, breathing problems, jaundice, etc.)? If yes, Please describe problems right after birth:
  - At how many days was your baby discharged home?
  - After the baby was born, how did he/she feed? If other, Please describe how he/she fed:
    - Breast
    - Bottle
    - Other
  - Did the baby have any feeding difficulties? If yes, Please describe any feeding difficulties:
  - Was your child born with any birth defects? (i.e. club foot, cleft lip and/or cleft palate, heart defects, extra fingers, etc.)? If yes, Please describe birth defects:
- Medical History. Now we will ask questions about your child's health history over their lifetime. Please answer the following Yes/No questions about possible tests/procedures/etc. that your child may have had. If your child has had one of these, please provide more detail in the box below (When? Why? Where? Results?).
  - Had a formal eye examination with ophthalmology?
  - Had a formal hearing examination?
  - Been hospitalized overnight?
  - Had surgery?
  - Currently taking any medications?
  - Been tested for or diagnosed with allergies/sensitivities?
  - Immunizations up to date?
- Does your child have any significant problems with any of the following?
  - Unusual weight gain or loss
    - Weight gain or loss?
    - Did weight gain ONLY occur while your child was taking a medication known to cause weight gain?
    - How much weight was gained?
    - Did weight loss ONLY occur while your child was taking a medication known to cause weight loss?
    - How much weight was lost?
  - Eye/Vision (e.g., glasses, strabismus, astigmatism)
    - Select any that apply:
      - Near-sighted (needs glasses for distance)
      - Far-sighted (needs glasses for reading)
      - Astigmatism
      - Strabismus (lazy/wandering eye)
      - Other
    - Did the strabismus require surgery to correct?
    - Please describe other visual problems here:
  - Hearing
    - Please describe hearing problem(s) here:
  - Ears/Nose/Mouth/Throat (e.g., infection, nose bleeds, dysphagia)
    - Select any that apply:
      - Recurrent ear infection
      - Recurrent nose bleed (epistaxis)
      - Tonsilitis (infection of tonsils)
      - Adenoiditis (infection of adenoids)
      - Dysphagia (trouble swallowing)
      - Other
    - Did any of the following require surgery to correct?
      - Ear infections (myringotomy/tube placement)
      - Nose bleeds (cauterization)
      - Tonsilitis (tonsilectomy)
      - Adenoiditis (Adenoidectomy)
    - Please describe other ear/nose/mouth/throat problems here:
  - Teeth (e.g., small, crowded, or misaligned teeth; prone to cavities)
    - Select any that apply:
      - Abnormally small teeth (microdontia)
      - Too many teeth (hyperdontia)
      - Too few teeth (hypodontia)
      - Overlapping or misaligned teeth (dental crowding)
      - Soft enamel or prone to cavities
      - Other
    - Please describe other tooth problems here:
  - Lungs/Breathing (e.g., asthma, recurrent infection, etc.)
    - Select any that apply:
      - Asthma
      - Frequent/recurrent respiratory infection
      - Other
    - Please describe other respiratory problems here:
  - Heart/Veins/Arteries/Circulation (e.g., heart defects, murmurs, high blood pressure)
    - Select any that apply:
      - Heart murmur
      - Congenital heart defect (any)
      - Syncope (fainting/passing out)
      - High blood pressure (hypertension)
      - Low blood pressure (hypotension)
      - Other
    - Please specify the congenital heart defect:
      - Aortic Valvar Stenosis (AS)
      - Aortic valve regurgitation
      - Atrial Septal Defect (ASD)
      - Atrioventricular Septal Defect (or AV Canal Defect)
      - Coarctation of the Aorta
      - Ebstein's Anomaly
      - Hypoplastic Left Heart Syndrome (HLHS)
      - Interrupted Aortic Arch/Ventricular Septic Defect
      - Mitral valve regurgitation
      - Patent Ductus Arteriosus (PDA)
      - Pulmonary Atresia (PA)
      - Pulmonary Valvar Stenosis (PS)
      - Pulmonary valve regurgitation
      - Single Ventricle Anomalies
      - Tetralogy of Fallot
      - Total Anomalous Pulmonary Venous Return
      - Transposition of the Great Arteries
      - Tricuspid Atresia
      - Tricuspid valve regurgitation
      - Truncus Arteriosus
      - Vascular Rings
      - Ventricular Septal Defect (VSD)
      - Other (specify below)
      - Unknown/unsure
    - Please specify the congenital heart defect:
    - Did the congenital heart defect require surgery to correct?
    - At what age was the surgery performed?
    - Please provide any additional details about the heart surgery. (Were there any complications? Were additional surgeries required? If so, at what ages?)
    - Please describe other cardiovascular problems here:
  - Stomach/Intestines/Bowels (e.g., constipation, nausea, reflux)
    - Select any that apply:
      - Constipation
      - Diarrhea
      - Nausea
      - Abdominal pain
      - Reflux (G.E.R.D.)
      - Inflammatory bowel disease (Crohn's, ulcerative colitis)
      - Irritable bowel syndrome (IBS)
      - Peptic ulcers
      - Other
    - Please describe other stomach, intestine, or bowel problems here:
  - Kidney/Bladder/Genitals (e.g., bed-wetting; genital abnormalities)
    - Select any that apply:
      - Abnormal genitalia
      - Enuresis (involuntary urination)
      - Other
    - Please describe the genital abnormality and whether surgery was required:
    - Enuresis (involuntary urination) occurs:
      - While awake
      - While asleep (bed-wetting)
      - Both awake and asleep
    - Age enuresis stopped (or select 'on-going' if it still occurs)
    - Please describe other kidney, bladder, or genital problems here:
  - Bones/Muscles/Joints (e.g., joint stiffness; scoliosis; musculoskeletal abnormalities)
    - Select any that apply:
      - Joint Pain
      - Joint Stiffness
      - Joint laxity/hypermobility ('double-jointed')
      - Scoliosis
      - Chest wall deformity
      - Hand/foot abnormality
      - Other
    - Please describe the chest wall deformity:
    - Please describe the hand/foot abnormality:
    - Please describe other bone, muscle, or joint problems here:
  - Skin/Hair/Nails (e.g., dermatitis, chronic acne)
    - Select any that apply:
      - Dry skin (xerosis)
      - Atopic dermatitis (eczema)
      - Psoriasis
      - Keratosis pilaris
      - Chronic acne
      - Abnormal toe/fingernails
      - Other
    - Please describe the toenail or fingernail abnormality:
    - Please describe other skin, hair, or nail problems here:
  - Easy bruising/bleeding or poor wound healing
    - Please describe the problem(s) related to easy bruising/bleeding or poor wound healing:
  - Headaches
    - Select all that apply:
      - During headaches, light or noise bothers him/her a lot more than usual
      - During headaches, he/she feels sick to their stomach
      - The headache pain is sometimes on only one side of their head
      - The headache pain sometimes throbs or comes in pulses
      - The headaches make it hard to do school work or other things they want to do
      - They have seen a doctor for their headaches
      - A doctor has said he/she has migraines
      - He/she has been treated for migraines
    - Other notes/details about headaches (optional):
  - Seizures
    - Select all that apply:
      - He/she has been diagnosed with epilepsy
      - He/she has been treated for epilepsy, convulsions, or seizures
      - Seizures are effectively treated with medication
      - A cause was identified for the seizures
      - Seizures ONLY occurred during a fever
    - What was identified as the cause of the seizure(s)
    - How many seizures has he/she had?
    - Age at first seizure:
    - Age at most recent seizure:
  - Loss of balance or coordination
    - Please describe the loss of balance or coordination problems:
  - Sleep disturbances/Problems
    - Select all that apply:
      - Sleep apnea
      - Difficulty falling asleep
      - Difficulty staying asleep
      - Sleep walking
      - Night terrors
      - Other
    - Please describe other sleep problems:
  - Fatigue
    - Please briefly describe the problems with fatigue here:
  - Growth/Feeding
    - Select all that apply:
      - Feeding problems during infancy
      - Feeding problems beyond infancy
      - Failure to thrive (slowed growth) during infancy
      - Failure to thrive (slowed growth) beyond infancy
      - Other
    - Please describe other problems with growth or feeding:
  - Heat or cold intolerance
    - Please describe the problems with heat or cold intolerance:
  - Delays or problems with puberty
    - Select any that apply:
      - Delayed puberty
      - Early (precocious) puberty
      - Other
    - Please describe the delays or problems with puberty:
  - Hormones/Endocrine (i.e. diabetes, thyroid problems)
    - Select any that apply:
      - Pre-diabetes (impaired glucose tolerance)
      - Type 1 diabetes (insulin dependent)
      - Type 2 diabetes (non-insulin dependent)
      - Hyperthyroidism (overactive thyroid)
      - Hypothyroidism (underactive thyroid)
      - Other
    - Please describe other hormone/endocrine problems here:
  - Serious Head Injury (that resulted in loss of consciousness)
    - At what age did it occur? (if more than one, age at first head injury)
    - How long did he/she lose consciousness for? (if more than one, refer to first head injury)
    - Please describe the head injury (what happened, was brain imaging performed, if so, what were the results). If more than one head injury occurred, please also include the ages when they occurred and how long he/she lost consciousness for.
  - Other significant medical problems not covered?
    - Please describe any other significant medical problems your child has experienced here:
- Diet/Feeding History
  - Restrictive eating? (Is he/she a particularly picky eater?)
    - Please select his/her preferred foods from the list: (foods he/she easily eats)
      - carbohydrates (breads, pastas, potatoes, etc.)
      - fruits (berries, apples, bananas, etc.)
      - leafy greens (spinach, broccoli, lettuce, etc.)
      - other vegetables (carrots, peas, squash, etc.)
      - dairy (milk, yogurt, cheese, etc.)
      - meats (beef, chicken, pork, etc.)
      - seafood (fish, crab, shrimp, etc.)
      - other proteins (eggs, nuts, etc.)
    - Please select his/her avoided foods from the list: (foods he/she refuses to eat)
      - carbohydrates (breads, pastas, potatoes, etc.)
      - fruits (berries, apples, bananas, etc.)
      - leafy greens (spinach, broccoli, lettuce, etc.)
      - other vegetables (carrots, peas, squash, etc.)
      - dairy (milk, yogurt, cheese, etc.)
      - meats (beef, chicken, pork, etc.)
      - seafood (fish, crab, shrimp, etc.)
      - other proteins (eggs, nuts, etc.)
  - Please describe any problems with your child's diet or feeding (either current or past problems):
- Parental Height and Weight
  - Biological father's height:
  - Biological father's weight:
  - Biological mother's height:
  - Biological mother's weight:
- Early Development
  - WHEN did you or your doctor first become concerned about your child's development?
  - If there were any concerns about your child's development, HOW were these concerns noticed?
- How old was your child when he/she began:
  - Rolling over?
  - Sitting alone?
  - Pulling to stand?
  - Crawling?
  - Cruising?
  - Walking alone?
  - First word?
  - First two-word phrases? (e.g. 'cat go', 'more juice'. Don't include words like 'bye-bye' or 'thank you' that are often used together)
  - First sentences?
  - Toilet trained?
  - Has your child lost any skills that he/she previously mastered (regression)?
    - Please describe any regression:
- School Information
  - Does your child currently attend school or day care?
    - What grade is he/she in currently?
  - Does your child attend special classes or need special help?
    - Please select the subject areas where your child requires special help (select all that apply):
      - Math
      - Reading
      - Writing
    - Please describe special classes your child attends or special help he/she needs (For example, what accommodations does he/she require? Is he/she in an inclusion class or self-contained class?):
    - If not already provided, please upload a copy of your child's most recent IEP (if possible):
- Has he/she ever received:
  - Physical Therapy
  - Occupational Therapy
  - Speech Therapy
  - Psychological Therapy/Counseling
  - How often does/did your child receive physical therapy?
  - How often does/did your child receive occupational therapy?
  - How often does/did your child receive speech therapy?
  - How often does/did your child receive psychological therapy?
- -
  - Does your child have any behavioral problems?
    - Please describe any behavioral problems:
  - Has a doctor ever diagnosed your child with any of the following learning problems? (Select all that apply)
    - Auditory processing disorder
    - Dyscalculia
    - Dyslexia
    - Dysphasia/Aphasia
    - Global developmental delay
    - Language (receptive) delay (problems understanding language)
    - Learning disability in math
    - Learning disability in reading
    - Mental retardation / intellectual disability
    - Non-verbal learning disability
    - Short term memory problems
    - Speech (expressive) delay (problems getting words out)
    - Verbal apraxia/dyspraxia
    - Visual processing deficits
    - Writing disability
    - None
    - Unsure
    - At what age was your child first diagnosed with the above learning problem(s)?
  - Has a doctor diagnosed your child with Autism?
    - At what age was your child diagnosed with Autism?
  - Has a doctor diagnosed your child with ADD or ADHD? (attention deficit hyperactivity disorder)
    - At what age was your child diagnosed with ADD/ADHD?
    - ADD/ADHD subtype:
      - Inattentive type
      - Hyperactive type
      - Combined type
      - Unsure/unknown
  - Has your child been diagnosed with the following psychiatric disorders (select any that apply):
    - Anxiety disorder
    - Bipolar / manic depression
    - Conduct disorder
    - Depression
    - Obsessive Compulsive Disorder
    - Oppositional defiant disorder
    - Panic attacks
    - Schizophrenia / schizoaffective
    - Other psychiatric disorder (not listed)
    - Unsure
    - None
    - At what age was your child first diagnosed with an anxiety disorder?
    - At what age was your child first diagnosed with bipolar disorder/manic depression?
    - At what age was your child first diagnosed with Conduct Disorder?
    - At what age was your child first diagnosed with depression?
    - At what age was your child first diagnosed with Obsessive Compulsive Disorder?
    - At what age was your child first diagnosed with Oppositional Defiant Disorder?
    - At what age was your child first diagnosed with Panic Attacks?
    - At what age was your child first diagnosed with Schizophrenia or Schizoaffective Disorder?
    - Please describe the other psychiatric disorder(s) and at what age they were diagnosed?
  - Do you feel that your child's language skills are where they should be for your child's age?
    - Please describe your child's language skills for his/her age:
  - Has your child ever had IQ testing or a formal developmental assessment?
    - When did your child receive IQ testing or a formal development assessment? And what were the results?
  - If not already provided, please upload a copy of the results from the most recent evaluation (if possible):

***4. Lifetime Psychiatric Medication History Survey***

Psychiatric Medication Histories are queried using a caregiver report survey. A list of commonly prescribed psychiatric medications is provided which includes brand and chemical names and can be sorted by indications for which they are commonly prescribed (see **Table S1**). Caregivers enter the name of each medication prescribed on a separate line, and are then asked to provide information on dose, duration, indication, perceived effectiveness, and observed side effects for each medication. There are open text boxes at the end of the survey where caregivers can include any additional relevant information.

***Survey Questions:***

- Has your child, either now or in the past, taken any prescription medications related to symptoms of ADHD, anxiety, depression, mood problems, schizophrenia, seizures/epilepsy, sleep disorders, or any other psychiatric problems?
- For reference, we have included a list of the most common psychiatric medications in alphabetical order. You may show the list by selecting one of the options on the right.
  - Show complete list
  - Show ADHD medications
  - Show anxiety medications
  - Show depression medications
  - Show schizophrenia medications
  - Show seizure medications/mood stabilizers (bipolar)
  - Show sleep medications
  - Hide list
  - [See **Table S1** for list of medications]
- Please list each psychiatric medication trial below. *Note about medication trials: If your child was prescribed the same medication at two different ages AND there was a period of 1 year or greater in between trials, please enter these as two separate medication trials below. If the medication was discontinued for less than one year in between trials, please consider this as one trial below.*
  - [Can add up to 15 medications]
- Please answer the following questions about [med1].
  - At what age was he/she first prescribed [med1]?
  - Prescribed for daily use or "as needed"?
    - Daily Use
    - As Needed (PRN)
  - For what indication (symptoms/diagnosis) was it prescribed?
    - ADHD
    - Agitation/aggression
    - Anxiety
    - Attention/concentration problems
    - Depression/sadness
    - Impulsivity
    - Mood problems (mania, bipolar)
    - Schizophrenia (psychosis)
    - Seizures/convulsions
    - Sleep problems (insomnia)
    - Other
    - Unknown
  - For other indications, please describe here:
  - Is he/she still taking [med1]?
    - At what age was it discontinued?
    - For what reason was it discontinued?
      - No Response/Poor Response (did not help symptoms)
      - Stopped working (symptoms came back)
      - Side effects/allergic reaction
      - Symptoms remitted (no longer needed)
      - Other
  - Have there been any side effects of the medication?
    - Please describe any side effects experienced and/or the reason why the medication was discontinued. (what happened; did side effects occur at higher dosages but not lower; were they immediate or delayed; did it require hospitalization)
  - On days when the medication is taken, what is the usual daily dose? (if no longer taking the medication, what was the usual daily dose)?
    - What is the current daily dose (in mg)?
    - What was the highest daily dose (in mg) taken for a period of six or more weeks? * Or check the box below if the medication was not taken for ≥ 6 weeks.
      - Check here if taken for less than 6 weeks:
    - What was the highest daily dose (in mg) taken for any length of time:
  - In your opinion, how effective was this medication at treating your child's symptoms?
    - Not effective at all
    - Minimally effective
    - Moderately effective
    - Extremely effective
- Has your child ever experienced an allergic reaction or serious side effects from ANY medication (including antibiotics and other non-psychiatric medications)? If yes, please list the medication(s) and briefly describe what happened: [OPEN TEXT FIELD]
- Is there anything else you think would be important for us to know? [OPEN TEXT FIELD]

***Table S1: List of Commonly Prescribed Psychiatric Medications***

| **Brand** | **Chemical** |
| --- | --- |
| Adderall | Amphetamine/dextroamphetamine |
| Strattera | Atomoxetine |
| Kapvay | Clonidine |
| Focalin | Dexmethylphenidate |
| Dexedrine | Dextroamphetamine |
| Tenex (Intuniv) | Guanfacine |
| Vyvanse | Lisdexamfetamine |
| Ritalin (Concerta, Dayrana, Metadate, Methylin) | Methylphenidate |
| Azstarys | Serdexmethylphenidate/dexmethylphenidate |
| Qelbree | Viloxazine |
| Elavil | Amitriptyline |
| Wellbutrin | Bupropion |
| Celexa | Citalopram |
| Anafranil | Clomipramine |
| Norpramin | Desipramine |
| Pristiq | Desvenlafaxine |
| Silenor | Doxepin |
| Cymbalta | Duloxetine |
| Lexapro | Escitalopram |
| Prozac | Fluoxetine |
| Luvox | Fluvoxamine |
| Fetzima | Levomilnacipran |
| Remeron | Mirtazapine |
| Paxil | Paroxetine |
| Zoloft | Sertraline |
| Desyrel | Trazodone |
| Effexor | Venlafaxine |
| Xanax | Alprazolam |
| Lexotan | Bromazepam |
| Librium | Chlordiazepoxide |
| Klonopin | Clonazepam |
| Valium | Diazepam |
| Hydroxyzine | Hydroxyzine |
| Ativan | Lorazepam |
| Serax | Oxazepam |
| Halcion | Triazolam |
| Abilify | Aripiprazole |
| Vraylar | Cariprazine |
| Thorazine | Chlorpromazine |
| Clozaril | Clozapine |
| Prolixin | Fluphenazine |
| Haldol | Haloperidol |
| Loxitane | Loxapine |
| Latuda | Lurasidone |
| Serentil | Mesoridazine |
| Moban | Molindone |
| Zyprexa | Olanzapine |
| Invega | Paliperidone |
| Trilafon | Perphenazine |
| Orap | Pimozide |
| Sparine | Promazine |
| Seroquel | Quetiapine |
| Risperdal | Risperidone |
| Mellaril | Thioridazine |
| Stelazine | Trifluoperazine |
| Geodon | Ziprasidone |
| Tegretol | Carbamazepine |
| Neurontin | Gabapentin |
| Vimpat | Lacosamide |
| Lamictal | Lamotrigine |
| Lithobid (Eskalith) | Lithium |
| Trileptal | Oxcarbazepine |
| Solfoton | Phenobarbital |
| Dilantin | Phenytoin |
| Topimax | Topiramate |
| Depakote (Devalproate, Depakene) | Valproic Acid |
| Quviviq | Daridorexant |
| Lunesta | Eszopiclone |
| Dayvigo | Lemborexant |
| Melatonin | Melatonin |
| Rozerem | Ramelteon |
| Belsomra | Suvorexant |
| Restoril | Temazepam |
| Sonata | Zaleplon |
| Ambien | Zolpidem |
